# Neuropsychiatric symptoms are associated with dementia in Parkinson’s disease but not predictive of it

**DOI:** 10.1101/2020.09.01.20186312

**Authors:** Kyla-Louise Horne, Michael R. MacAskill, Daniel J. Myall, Leslie Livingston, Sophie Grenfell, Maddie J. Pascoe, Bob Young, Reza Shoorangiz, Tracy R. Melzer, Toni L. Pitcher, Tim J. Anderson, John C. Dalrymple-Alford

**Affiliations:** New Zealand Brain Research Institute, Christchurch, New Zealand; Department of Medicine, University of Otago, Christchurch, New Zealand; Brain Research New Zealand Rangahau Roro Aotearoa Centre of Research Excellence; School of Psychology, Speech and Hearing, University of Canterbury, New Zealand; Department of Electrical and Computer Engineering, University of Canterbury, New Zealand; Department of Neurology, Christchurch Hospital, Christchurch, New Zealand

**Keywords:** Neuropsychiatric symptoms, PDD risk, Prediction, Parkinson’s dementia, Longitudinal

## Abstract

**Background:** Neuropsychiatric symptoms in Parkinson’s disease (PD) may increase dementia (PDD) risk. The predictive value of these symptoms, however, has not been compared to clinical and demographic predictors of future PDD.

**Methods:** 325 PD participants completed baseline neuropsychiatric and MDS-Task Force Level II assessments. Of these, 195 non-demented individuals were followed-up over a four-year period to detect conversion to PDD; 51 developed PDD. ROC analysis tested associations between baseline neuropsychiatric symptoms and conversion to PDD. The probability of developing PDD was also modelled as a function of neuropsychiatric inventory (NPI) total score, PD Questionnaire (PDQ) hallucinations, PDQ anxiety and contrasted to cognitive ability, age and motor function. Leave-one-out information criterion was used to evaluate which models provided useful information when predicting future PDD.

**Results:** The PDD group experienced greater levels of neuropsychiatric symptoms compared to the PD-MCI and PD-N groups at baseline. Few differences were found between the PD-MCI and PD-N groups. Five neuropsychiatric measures were significantly, but weakly, associated with future PDD. The strongest was NPI total score: AUC=0.66 [0.55-0.76]. There was, however, no evidence that it contained useful out-of-sample predictive information of future PDD (delta ELPD=1.6 (SD 2.4)); Similar results held for PDQ hallucinations and PDQ anxiety. In contrast, cognitive ability (delta ELPD=35 (SD 8)) and age (delta ELPD=11 (SD 5)) provided useful predictive information of future PDD.

**Conclusions:** Cognitive ability and age strongly out-performed neuropsychiatric measures as markers of developing PDD within four years. Therefore, neuropsychiatric symptoms do not appear to be useful markers of PDD risk.

## Introduction

The presence of neuropsychiatric symptoms in Parkinson’s disease (PD) may represent early markers of progression to dementia (PDD). There is evidence that visual hallucinations (OR=3-10), illusions (OR=8), thought disorders^[1, 2]^, depression (OR=2.3)^[1]^ and apathy (HR=7.2; OR=1.1)^[1, 3]^ are associated with future PDD.

Neuropsychiatric symptoms may, however, provide minimal predictive information compared to older age and cognitive decline, both established risk factors for PDD^[4]^. The former includes associations between older age per se (OR=2.3; HR=1.1) and older age at PD onset (OR=2.2) with risk of PDD^[4, 5]^. The latter factor includes baseline PD-MCI status (OR=10.2; HR=3.5-11.3), subjective cognitive complaints (OR=2.6) and lower cognitive ability (OR=0.8-10.1) ^[1, 4, 5]^. Motor features, such as gait impairment (OR=1.2), falls (OR=3.0), freezing (OR=2.6), UPDRS III (HR=1.0) and mixed tremor/akinetic subtype (OR=3.3) are also associated with PDD risk^[1, 2, 4]^. Awareness of the importance of neuropsychiatric features to the risk of PDD therefore requires an understanding of their contribution compared to age, cognition and clinical motor features. These comparisons have not been reported previously.

Using a well-characterised, longitudinal cohort of PD participants who have been assessed using Level II Movement Disorder Society-Task Force (MDS-TF) PD-MCI criteria, we (1) compared neuropsychiatric symptoms across three cognitive status groups (i.e. PD-N, PD-MCI and PDD). Then we (2) examined the association of neuropsychiatric symptoms and conversion to PDD in a large sample of non-demented PD participants who were followed up over a four-year period. We then (3) determined the clinical out-of-sample predictive utility of a subset of neuropsychiatric symptoms compared to age, cognitive ability and motor scores.

## Methods

**Participants**. A baseline convenience sample of 325 PD participants (178 PD-N, 107 PD-MCI and 40 PDD) completed MDS-TF Level II neuropsychological assessments^[6]^, along with neuropsychiatric and clinical measures (Figure 1). There were 195 PD participants who were non-demented at baseline (i.e. PD-N and PD-MCI) and their conversion to PDD within a 4-year window could be assessed. Individuals were classified as a converter if they had an assessment within 4.5 years post-baseline where they were classified as PDD; otherwise they were classified as a non-converter if they had an assessment 3.5 years or later post-baseline where they were classified as PD-N or PD-MCI. At study entry, two PDD participants did not receive the Geriatric Depression Scale (GDS) and one of those also did not receive the Parkinson’s Disease Questionnaire (PDQ). Two PD-N, two PD-MCI and three PDD participants did not have their motor symptoms evaluated at baseline. The comprehensive neuropsychological assessments, PD diagnostic procedure and exclusion criteria are detailed in Wood et al. (2016). The mean duration of motor symptoms at study entry was 7.7 years (SD 5.4 years). All participants took their usual medications on the day of testing. The study was approved by the Upper South B Regional Ethics Committee of the New Zealand Ministry of Health. At the beginning of each neuropsychological assessment, participants each read and sign a consent form after discussing any questions or concerns they may have. All participants gave informed written consent.

**Figure 1.**
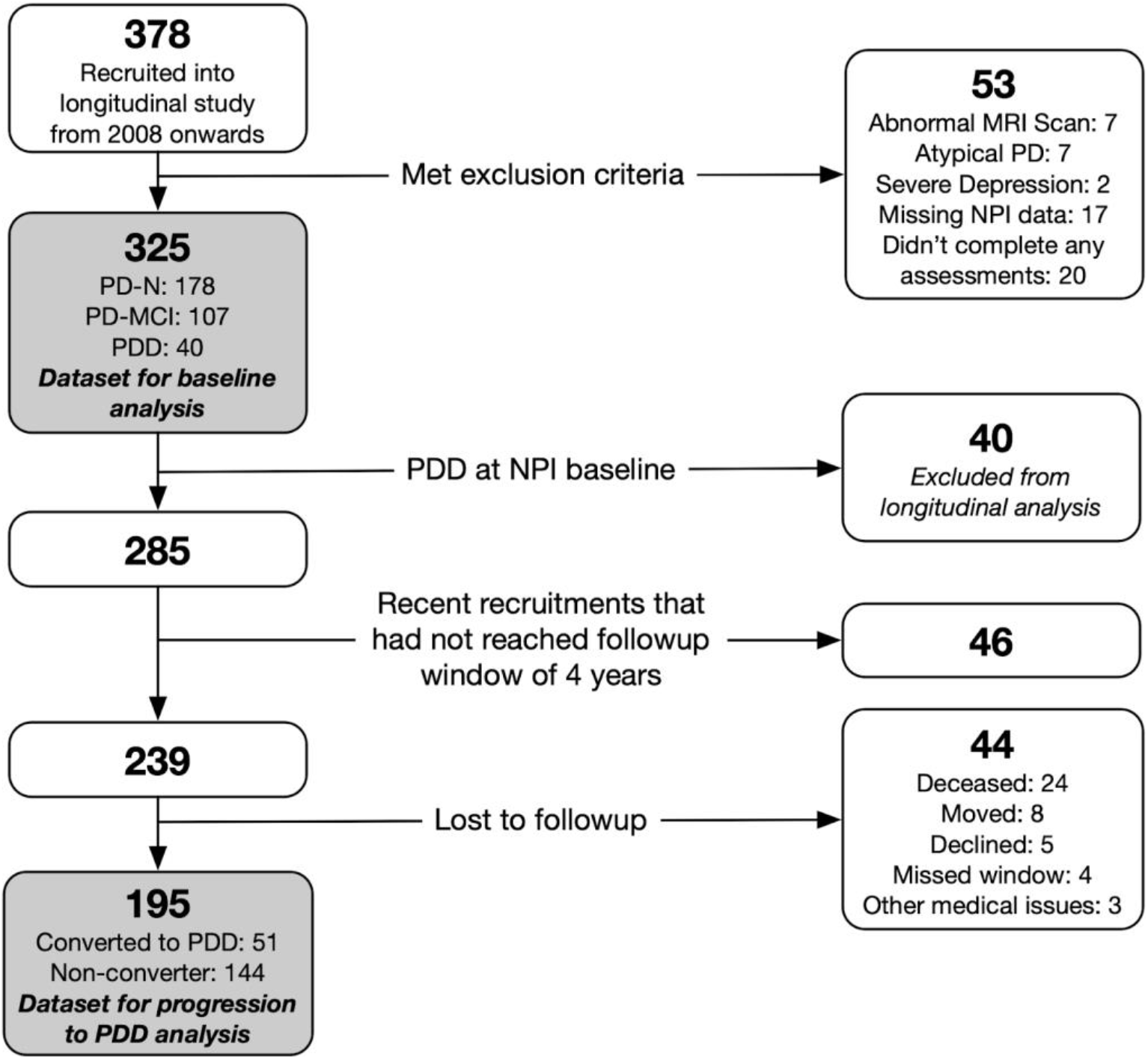
Participant recruitment, exclusions and total followed over four years. NPI = Neuropsychiatric Inventory; PD = Parkinson’s disease; PDD = Participants who met Level II criteria for Parkinson’s disease with dementia; PD-MCI = Participants who met Level II criteria for Parkinson’s disease with mild cognitive impairment; PD-N = The remaining Participants who did not meet criteria for PDD or PD-MCI were classified as having normal cognition. *Assessments were conducted at baseline and then again every one to two years subsequently; participants with dementia were not followed further*.

**Determination of cognitive status**. PD participants were classified as PDD using established MDS-TF PDD criteria^[8]^. The remaining participants were classified at baseline as either PD-MCI (using a criterion of at least two test impairments at least −1.5SD below norms in a single cognitive domain))^[7]^, consistent with MDS-TF Level II PD-MCI guidelines^[6]^, or PD-N (not meeting either PD-MCI or PDD criteria).

**Neuropsychiatric assessments**. These were as follows: (i) The Neuropsychiatric Inventory (NPI), reported by a PD participant’s significant other (SO),^[9, 10]^; (ii) 15-item GDS (Geriatric Depression Scale)^[11, 12]^; (iii) the hallucination item, distressing dreams item and the four-items from the emotional well-being sub-section of the PDQ (Parkinson’s Disease Questionnaire)^[13]^; and (iv) two items indicative of hallucinations and depression extracted from Part I of the Unified PD Rating Scale (UPDRS)^[14, 15]^. UPDRS Part III data were converted to MDS-UPDRS scores ^[16]^.

**Analysis**. This was conducted in three parts, in the R statistical environment (v.3.6.3)^[17]^. First, baseline demographic and neuropsychological profiles of the three PD groups were compared using ANOVA followed by Tukey post hoc tests. The baseline neuropsychiatric symptoms were then compared pairwise across the PD-N, PD-MCI and PDD groups using receiver operator characteristic (ROC) tests to determine the discrimination between baseline groups based on neuropsychiatric symptoms. The ROC analysis included boot-strapped confidence intervals (*pROC)*^[18]^. Second, we examined the association between baseline neuropsychiatric measures and conversion or not to PDD using ROC tests in the 195 non-demented PD participants. Lastly, we evaluated the clinical utility of the selected statistically significant neuropsychiatric measures identified by the ROC analysis along with other previously-proposed predictors (baseline cognitive ability, age and motor impairment) using Bayesian regression models (*brms* (v2.9.0)^[19]^. We applied the regression models to 192 non-demented participants (three participants were excluded as they were not administered the UPDRS at baseline). The null model consisted only of an intercept, this was used for comparison. Six models then examined each of six predictors separately: three neuropsychiatric measures from the ROC analysis, global cognitive score [expressed as an aggregate z score derived from averaging performance from measures conducted in four cognitive domains], age, and motor score. The dependent variable was binary: progression or non-progression to PDD. The leave-one-out information criterion (LOO-IC)^[20]^, which approximates the expected log pointwise predictive density [ELPD], was used to estimate the predictive accuracy of models. The difference in ELPD of the six models to the null model was calculated, and a delta ELDP of greater than twice the SE of the estimate was taken as evidence that a model was an improvement over the null model. Finally, a model, comprising all six measures, was updated to determine the conditional effects of each predictor.

## Results

### Group differences on baseline neuropsychiatric symptoms and cognitive status

Table 1 depicts the baseline demographic, neuropsychological and neuropsychiatric profiles of the three PD groups. Education was similar across cognitive groups. The PDD group was older, had more advanced PD, greater motor burden and of course showed increased everyday functional-impairments than the other PD groups at baseline. The PD-MCI group showed intermediate mean values compared to the PD-N and PDD groups, except symptom duration was similar between the two non-dementing groups.

**Table 1.** Demographics, neuropsychological and neuropsychiatric measures at study entry [mean (SD)]

|  | PD-N | PD-MCI | PDD |
| --- | --- | --- | --- |
| <u>Demographics</u> |  |  |  |
| Sample size (n) | 178 | 107 | 40 |
| Converted to PDD (n) | 8 | 43 | N/A |
| Sex, M:F | 115:63 | 81:26 | 30:10 |
| Age | 68 (8) | 71 (7)<sup>a</sup> | 74 (7)<sup>a,b</sup> |
| Education (y) | 13 (3) | 13 (3) | 13 (3) |
| <u>PD-Specific Measures</u> |  |  |  |
| Symptom duration (y) | 7 (5) | 8 (5) | 12 (7)<sup>a,b</sup> |
| Hoehn and Yahr stage | 2 (0.5) | 2 (0.7)<sup>a</sup> | 3 (0.7)<sup>a,b</sup> |
| UPDRS (Motor) | 29 (13) | 39 (15)<sup>a</sup> | 57 (19)<sup>a,b</sup> |
| <u>Activities of Daily Living</u> |  |  |  |
| CDR | 0.1 (0.2) | 0.4 (0.2)<sup>a</sup> | 1.3 (0.5)<sup>a,b</sup> |
| Reisberg IADL | 0.5 (0.5) | 0.8 (0.5)<sup>a</sup> | 2.1 (0.6)<sup>a,b</sup> |
| <u>Cognitive Assessments</u> |  |  |  |
| Premorbid IQ (WTAR) | 111 (8) | 108 (10)<sup>a</sup> | 107 (11)<sup>a</sup> |
| DRS-2 (AESS) | 12 (2) | 9 (3)<sup>a</sup> | 5 (3)<sup>a,b</sup> |
| ADAS-Cog | 6.6 (2.8) | 10.8 (3.9)<sup>a</sup> | 22.5 (8.6)<sup>a,b</sup> |
| MoCA | 27 (2) | 23 (3)<sup>a</sup> | 17 (4)<sup>a,b</sup> |
| Global Z score | 0.18 (0.42) | -0.84 (0.51) | -1.91 (0.48) |
| <u>NPI measures</u> |  |  |  |
| NPI Total score | 3.8 (6.8) | 4.3 (6.8) | 10.8 (8.2) |
| NPI Hallucinations | 0.1 (0.8) | 0.3 (1.2) | 1.3 (2.8) |
| NPI Depression | 0.7 (1.7) | 0.9 (2.0) | 1.7 (3.0) |
| NPI Anxiety | 1.0 (2.3) | 1.2 (2.0) | 1.9 (2.4) |
| NPI Aggression | 0.2 (1.3) | 0.2 (1.1) | 1.2 (2.1) |
| NPI Apathy | 0.7 (1.8) | 0.7 (1.7) | 2.1 (2.9) |
| NPI Disinhibition | 0.2 (1.0) | 0.1 (0.5) | 0.6 (1.9) |
| NPI Delusions | 0.2 (1.3) | 0.0 (0.1) | 0.8 (2.4) |
| NPI Euphoria | 0.0 (0.2) | 0.0 (0.3) | 0.0 (0.0) |
| NPI Irritability and Liability | 0.5 (1.7) | 0.4 (1.5) | 0.6 (1.6) |
| NPI Aberrant Motor Behaviour | 0.2 (1.1) | 0.4 (1.3) | 0.6 (1.7) |
| NPI Sleep and Night time Behaviour Disorders | 1.7 (3.0) | 2.0 (3.2) | 1.5 (2.7) |
| NPI Appetite and Eating Disorders | 0.5 (1.7) | 0.5 (1.8) | 0.7 (2.1) |
| <u>PDQ measures</u> |  |  |  |
| PDQ Hallucinations | 0.4 (0.8) | 0.6 (1.0) | 1.1 (1.3) |
| PDQ Depression | 0.7 (1.0) | 0.8 (1.0) | 1.3 (1.2) |
| PDQ Anxiety | 1.0 (1.0) | 1.1 (1.0) | 1.6 (1.2) |
| PDQ Aggression | 0.3 (0.6) | 0.4 (0.8) | 0.8 (1.0) |
| PDQ Isolated and Lonely | 0.4 (0.7) | 0.4 (0.8) | 1.0 (0.9) |
| PDQ Weepy and Tearful | 0.6 (0.9) | 0.6 (0.9) | 0.7 (0.7) |
| PDQ Worried about future | 1.1 (1.1) | 1.1 (1.2) | 1.1 (1.0) |
| PDQ Distressing dreams | 0.5 (0.9) | 0.6 (1.0) | 1.3 (1.4) |
| <u>UPDRS measures</u> |  |  |  |
| UPDRS Hallucinations | 0.3 (0.6) | 0.5 (0.9) | 1.3 (1.2) |
| UPDRS Depression | 0.5 (0.8) | 0.6 (0.9) | 1.2 (1.0) |
| <u>GDS</u> | 0.1 (0.4) | 0.1 (0.3) | 0.5 (0.6) |

*Abbreviations: PDD = Participants who met Level II criteria for Parkinson's disease with dementia; PD-MCI = Participants who met Level II criteria for Parkinson's disease with mild cognitive impairment; PD-N = The remaining Participants who did not meet criteria for PDD or PD-MCI were classified as having normal cognition; ADAS-Cog = Alzheimer's Dementia Assessment Scale-Cognitive; CDR = Clinical Dementia Rating; DRS-2 (AESS) = Dementia Rating Scale-2 (Age and Education Scaled Score); GDS = Geriatric Depression Score; Global Z score = mean derived from attention, executive function, visuospatial and episodic memory domains; IADL = Instrumental Activities of Daily Living; MoCA = Montreal Cognitive Assessment; NPI = Neuropsychiatric Inventory; PDQ = Parkinson's disease Questionnaire; UPDRS = Unified Parkinson's Disease Rating Scale; WTAR = Wechsler Test of Adult Reading for premorbid IQ.*
<sup>a</sup>Significantly different from PD-N, ANOVA followed by Tukey post hoc tests, $p < 0.05$ .
<sup>b</sup>Significantly different from PD-MCI, ANOVA followed by Tukey post hoc tests, $p < 0.05$ .

Figure 2 depicts the baseline AUC values for each neuropsychiatric symptom. Significantly greater levels of hallucinations, depression, anxiety, and aggression in PDD versus PD-N were present in multiple measures from the NPI, PDQ, UPDRS and GDS scales. Sub-components of the NPI and PDQ also indicated greater levels of apathy, loneliness, and distressing dreams in PDD versus PD-N. When PDD and PD-MCI groups were compared, a similar overall pattern was seen, but the AUC values were generally smaller; additionally, there was no evidence of a difference in anxiety scores, but there was evidence of a difference in the level of delusions between these groups. There were few significant differences between the PD-N and PD-MCI groups, with marginally higher levels of NPI and UPDRS hallucinations, NPI anxiety and NPI aberrant motor behaviour item scores in the PD-MCI group compared to the PD-N group.

**Figure 2.**
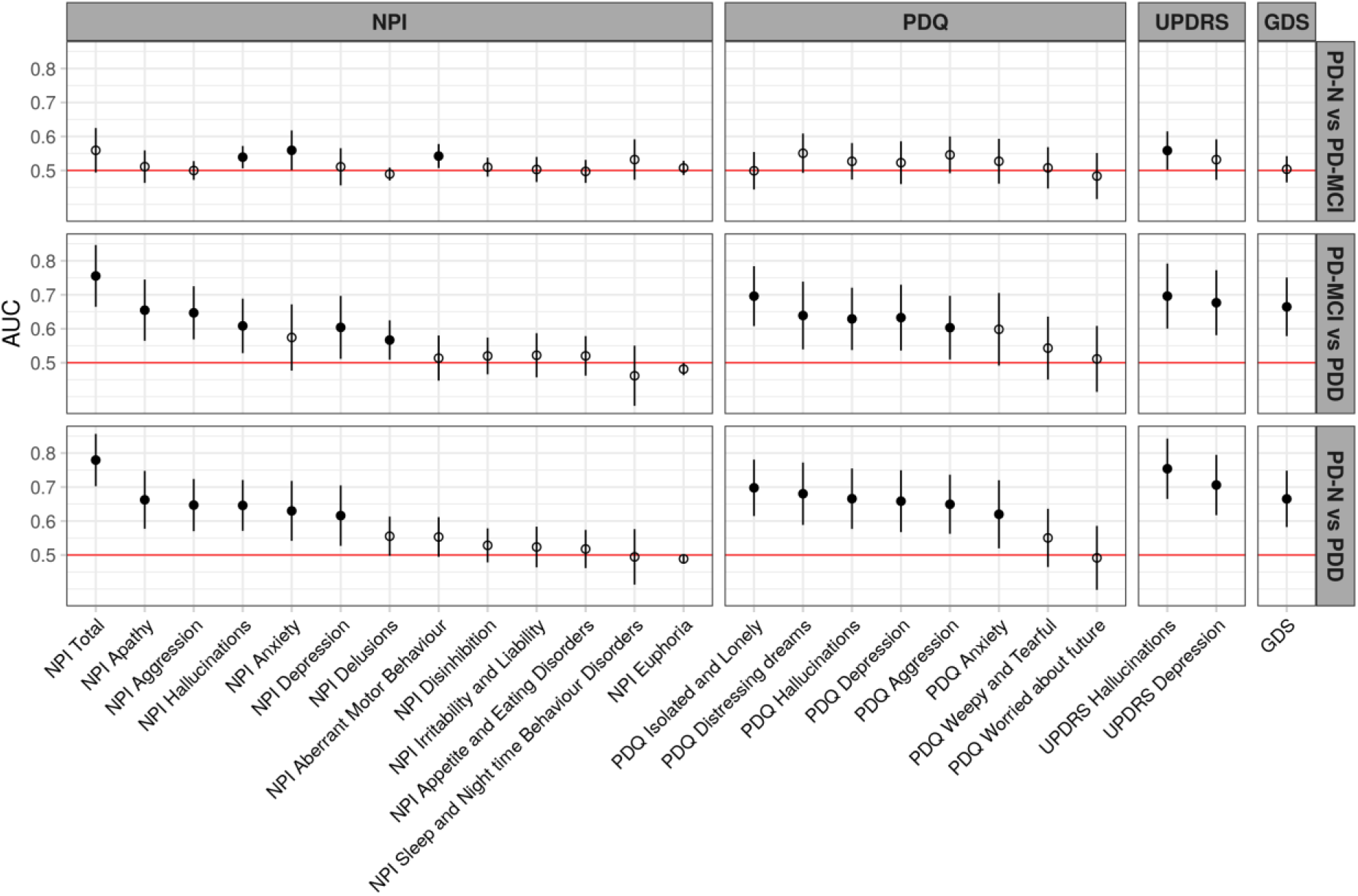
AUC values represent the probability of a classifier using individual behavioural symptom scores to rank a randomly chosen participant from one diagnostic group higher than a participant from another group. The dots represent the estimated AUC values, lines represent 95% CI. Filled dots indicate values significantly greater than 0.5 (chance performance). Items are from the NPI, PDQ, MDS-UPDRS, and GDS scales. Items within each scale are sorted from right to left by the AUC value for classifying PDD from PD-N.

### Baseline neuropsychiatric symptoms associated with progression to PDD within four years

In the non-demented PD participants, baseline neuropsychiatric symptoms were examined for differences between those who converted to PDD (n=51; 43 from PD-MCI and 8 from PD-N) and those who did not (n=144). Five neuropsychiatric symptoms had significant AUC values (Figure 3). NPI total score produced the largest but nonetheless weak association with conversion to PDD (AUC=0.66 [0.55-0.76]). The PDQ hallucinations item (AUC=0.63 [0.55-0.70]), NPI aberrant motor behaviour item (AUC=0.56 [0.51-0.62]), PDQ anxiety item (AUC=0.60 [0.51-0.69]) and NPI anxiety item (AUC=0.61 [0.53-0.70]) also had weak associations with conversion to PDD. No other neuropsychiatric measures were associated with conversion to PDD. We selected the NPI total score, PDQ hallucination and PDQ anxiety items for subsequent analysis. The NPI anxiety and aberrant motor behaviour items were not used, although they were both weakly associated with future PDD by themselves, because the NPI total score is a summation of all the NPI components.

**Figure 3.**
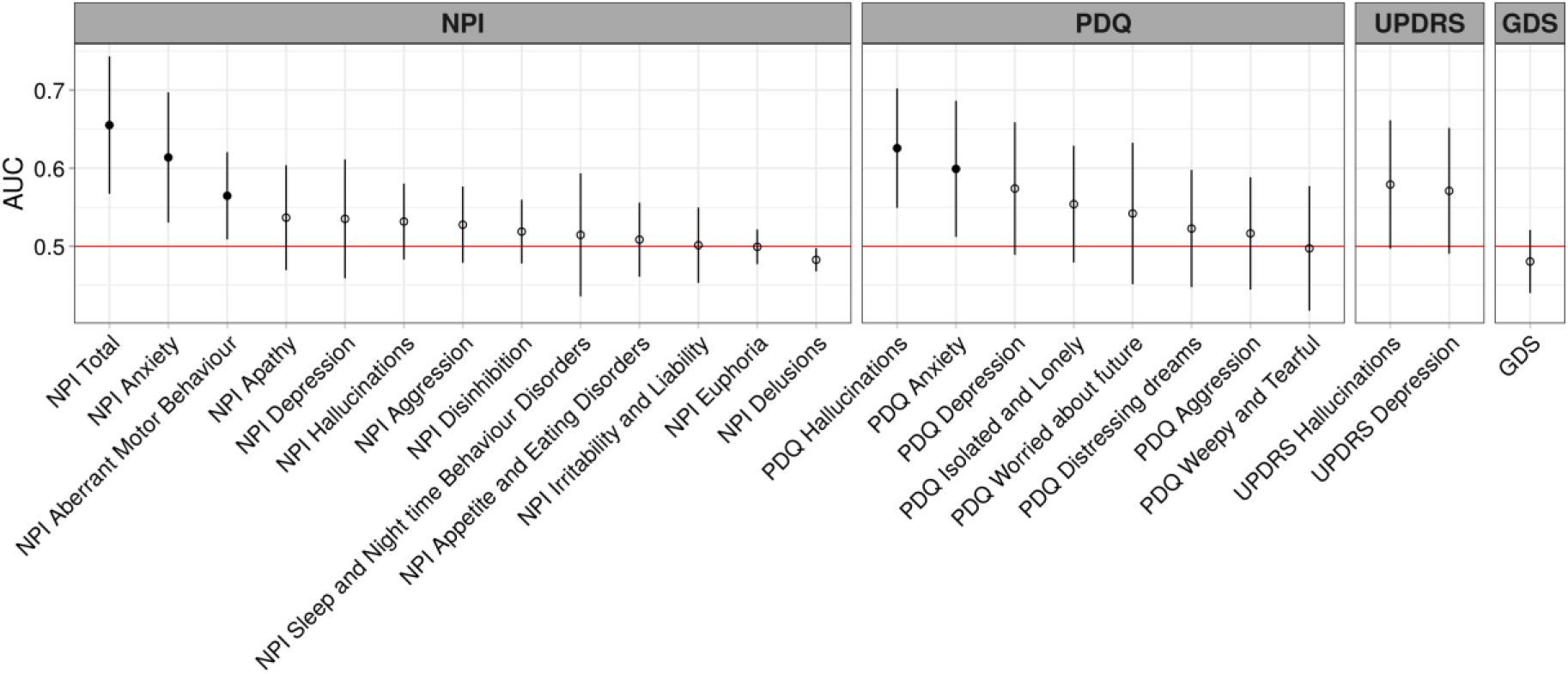
AUC values for conversion to PDD across neuropsychiatric measures. The dots represent AUC values, lines represent 95% CI and filled dots indicate values significantly greater than 0.5.

### Determining useful predictors of future progression to PDD

The improvement in the model fit compared to a null model with only the intercept for each of the three separate neuropsychiatric measure models was very weak (NPI total score, ELPD=1.6 (SD 2.4); hallucinations, delta ELPD=2.5 (SD 2.8); anxiety, delta ELPD=2.0 (SD 2.5)). That is, none of these neuropsychiatric measures provided reliably useful information to predict future dementia. In contrast, independent models containing global cognitive score only (delta ELPD=35 (SD 8)) and age only (delta ELPD=11 (SD 5)) had much stronger associations with future PDD. Motor function had minimal association with the probability of future PDD (delta ELPD=5 (SD 3)). For ease of comparison with previous studies, odds ratios were calculated for each of the three neuropsychiatric measures. At baseline, NPI total score, PDQ hallucinations and PDQ anxiety were all weakly associated with progression to PDD within four years (estimated odds ratio, NPI total score, 1.1 [1.0-1.1]; hallucinations, 1.6 [1.1, 2.3]; anxiety, 1.5, [1.1-2.1]).

The neuropsychiatric measures and previously-proposed predictors (age, cognitive ability and motor function) were then included in a single model with the conditional effects of developing PDD four years later shown in Figure 4. Along the *x*-axis of each of the six plots displayed in Figure 4 are the range of possible scores for each predictor, while the probability of PDD conversion is displayed on the *y*-axis. Figure 4a shows the conditional probability of PDD conversion given NPI total score. There is a large amount of variability across each of the samples for the NPI total score measure and a mean probability of progression to PDD of <30% across the possible range of scores. The same variability across the samples can be seen for both PDQ hallucinations (Figure 4b) and PDQ anxiety (Figure 4c), with mean probability of progression to PDD of <30% and <25% across the possible range of scores respectively. In contrast, global cognitive score (figure 4d) is a much better predictor of future PDD progression as the samples are more tightly clustered together and there is a probability of converting to PDD of >55% when the global cognitive score is worse than −1. In figure 4e, older age increases the probability of future PDD, however there is also an increased sample variability when age is greater than 80 years. In Figure 4f, however, although the UPDRS III score samples are tightly clustered together, the probability of converting to PDD is <10% across the possible range of scores. It therefore appears that UPDRS III score has essentially no predictive value. Overall, in the combined model of global cognitive score, age, NPI total score, PDQ hallucinations, PDQ anxiety and UPDRS motor score, only cognition and age contributed useful predictive information of conversion to PDD within four years.

**Figure 4:**
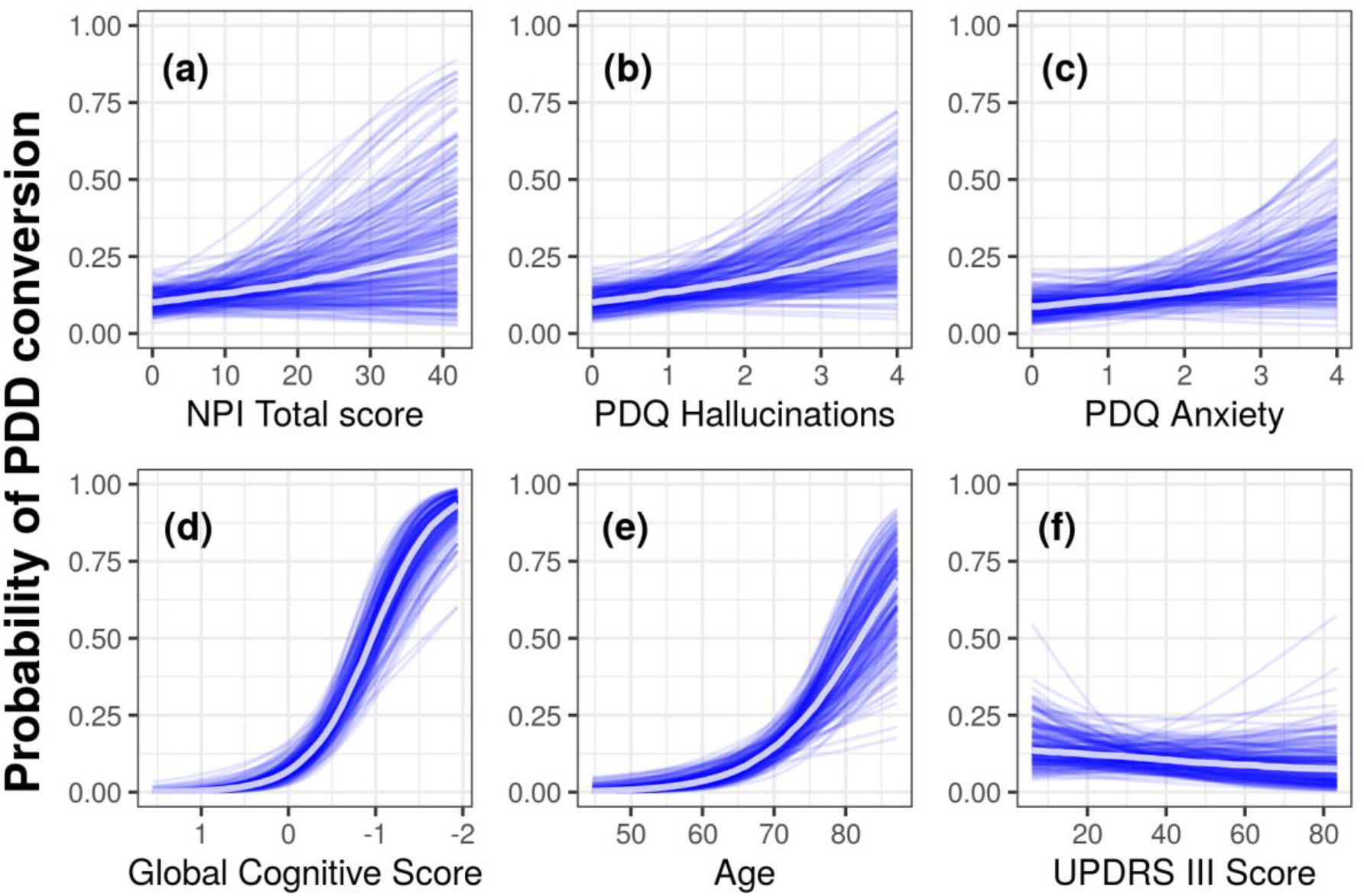
Conditional probability (with other variables held at their mean value) of conversion to PDD within four years as a function of (a) NPI Total Score, (b) PDQ hallucinations, (c) anxiety (d) Global cognitive score with scores worsening from left to right, (e) age, and (f) UPDRS III motor score. Each blue line represents one sample from the posterior, and the central white line represents the mean of those posterior samples. The mean values of each predictor was NPI Total Score = 4.2, PDQ Hallucinations = 0.5, PDQ Anxiety = 1.1, Global cognitive score = −0.2, age = 68.3, and UPDRS III motor score = 31.6.

## Discussion

At baseline, we found that PDD participants experienced greater neuropsychiatric symptom burden than did non-demented PD. Over the four-year follow-up period, of the neuropsychiatric measures, baseline NPI total score, PDQ hallucinations, NPI aberrant motor behaviour and the anxiety measures from the NPI and PDQ all had weak associations with PDD risk. However, when the NPI total score, PDQ hallucination and PDQ anxiety were examined in a Bayesian regression model and the out-of-sample predictive ability compared to age, cognitive ability and motor function, none of the neuropsychiatric symptoms were useful when predicting future PDD.

Only five neuropsychiatric symptoms were significantly associated with progression to PDD four years later. However, global cognitive ability and age, rather than neuropsychiatric symptoms, were better predictors of progression to PDD. Unlike previous studies, we did not find an association between baseline apathy measures and future dementia risk^[3, 21]^ or baseline depression measures and progression to PDD four-years later^[1]^. Nevertheless, we did find an association between total NPI score, hallucinations, anxiety and aberrant motor behaviour and progression to PDD. The relationship between future PDD and hallucinations has been reported by others^[1, 2, 22]^. These studies vary in sample size from 80-224 PD participants (followed for four-eight years) and all rated the presence of hallucinations using a single hallucination measure from the UPDRS, whereas we included three hallucination measures in this study (UPDRS, PDQ, and NPI). We found the odds ratio of developing PDD for participants who experienced hallucinations versus those who did not was 1.6 [1.1, 2.3]. This was much lower than previous studies (OR=3.1-10.2)^[1, 2]^. However, strengths of our study were the additional comparisons made to determine whether hallucinations provide useful information compared to previously-proposed risk factors for PDD, which have not been evaluated in previous studies. These suggested that while neuropsychiatric symptoms are common in non-demented PD^[23, 24]^ and have a negative impact on quality of life^[25, 26]^, they provide no predictive information about PDD development, at least over a four year period.

Surprisingly the severity of motor symptoms did not provide any useful predictive information. Many previous studies have found an association between a variety of PD motor symptoms, including freezing, gait impairment and falls, and increased PDD risk^[1, 2, 4]^. In this study however, motor impairment, as measured by the UPDRS III score, did not add any useful predictive value once age and cognitive ability were known. This finding is supported by two recent multinational studies which both found that when UPDRS III scores were accounted for, cognitive impairment independently increased PDD risk^[4, 27]^.

Only the NPI hallucinations, anxiety, aberrant motor behaviour and UPDRS hallucinations items differed significantly when comparing neuropsychiatric symptoms between PD-MCI and PD-N. Previous studies have found mixed results. Broeders et al. (2013) found that PD-MCI patients reported higher rates of depression and anxiety compared to PD-N, whereas other studies have found no differences in depression^[29, 30]^, hallucinations^[30]^ and NPI total scores^[29]^. These mixed findings could be due to differences in the tests employed to measure each of these symptoms. We used multiple tests to measure depression (NPI, PDQ, UPDRS and GDS) and anxiety (NPI and PDQ), whereas Broeders et al. (2013) used the Hospital Anxiety and Depression Scale (HADS). Both the GDS and HADS have clinical utility in detecting depression in PD^[31]^ but could be capturing different aspects of depression symptomatology. Variations in the tests employed to measure each neuropsychiatric symptom might therefore explain the differences between these studies.

At baseline, the PDD group presented with more severe hallucinations, depression, anxiety, apathy, loneliness, distressing dreams and aggression compared to PD-N. A similar overall pattern was seen when the PDD group was compared to PD-MCI. No difference in anxiety levels, however, was found between PDD and PD-MCI. These cross-sectional findings at baseline align with previous studies examining the prevalence of neuropsychiatric symptoms, with those with PDD having greater prevalence than those with PD-MCI and PD-N^[29, 30, 32]^. Thus, it is clear from this and other studies that the development of dementia in PD brings with it a greater burden of neuropsychiatric symptoms.

There were few participants who experienced certain neuropsychiatric symptoms, such as delusions (n=15), disinhibition (n=19) and euphoria (n=8) as measured by the NPI. This could be due to the scales lacking the sensitivity to detect these symptoms, or that they are uncommon in PD. Aside from the GDS, the other neuropsychiatric items used in this study were not specifically designed to be used as single measures, separate from the overall scales within which they sit. Thus, those items may not be as sensitive in determining the true prevalence of these symptoms in PD populations. The use of single items extracted from larger scales is a potential limitation of this study, although most other studies examining the relationship between hallucinations and future PDD have also used single measures (from the UPDRS)^[1, 2, 22]^. We recommend that future studies employ more specifically-targeted neuropsychiatric symptom scales (such as the Psychosis and Hallucinations Questionnaire^[33, 34]^ or HADS), to determine the true predictive value of each neuropsychiatric symptom.

In summary, we found that five neuropsychiatric symptoms present at baseline in non-demented PD patients were weakly associated with progression to PDD four-years later, NPI total score, PDQ hallucinations, PDQ anxiety, NPI anxiety and NPI aberrant motor behaviour. We also confirmed that participants with PDD experienced more neuropsychiatric symptoms than those with those with either PD-MCI or PD-N, with no difference in their prevalence between the two non-dementing groups. When NPI total score, PDQ hallucinations, NPI anxiety and motor function were examined alongside cognitive impairment and age, they did not have any useful predictive value, which suggests that they are not useful markers of future PDD risk but instead evolve in parallel with the disease process.

## Data Availability

The data and code used for this analysis are currently available from the authors upon request. We are in the process of producing a properly anonymised dataset along with analysis code which will be made available for download, likely from the Open Science Framework website, with a link back to this pre-print.

## Acknowledgments

We would like to acknowledge individuals who have collected data, including Saskia van Stockum, Charlotte Graham, Krysta Callander, Megan Livingstone and Beth Elias.We would also like to thank and acknowledge funding from the New Zealand Health Research Council, Brain Research New Zealand, Neurological Foundation of New Zealand, Canterbury Medical Research Foundation, and the New Zealand Brain Research Institute.Author roles:

1) Research project: A. Conception, B. Organization, C. Execution;

2) Statistical Analysis: A. Design, B. Execution, C. Review and Critique;

3) Manuscript: A. Writing of the first draft, B. Review and Critique.

**K-LH:** 1A, 1B, 1C, 2A, 2B, 2C, 3A, 3B; **MRM:** 1A, 1B, 2A, 2B, 2C, 3B; **DJM:** 1A, 2A, 2B, 2C, 3B; **LL:** 1B, 1C, 3B; **SG:** 1C, 3B; **MP:** 1C, 3B; **BY:** 1C, 3B; **RS:** 2B, 2C, 3B; **TRM:** 2C, 3B; **TLP:** 1B, 1C, 3B; **TJA:** 1A, 1B, 2C, 3B; **JCD-A:** 1A, 1B, 2A, 2C, 3B.

